# First Detection of Xylazine in Texas Wastewater and Its Association with Fentanyl Use

**DOI:** 10.1101/2024.11.19.24317580

**Authors:** Katherine M. Joseph, Dhvani Parikh, Qin Xuan, Feng Li, John Balliew, Kristina D. Mena, Fuqing Wu

## Abstract

The United States is dealing with the drug overdose crisis that has intensified over the past decade and compounded by the emergence of new threats particularly xylazine, a veterinary sedative increasingly found in illicit drug supplies. This study investigates the prevalence of xylazine in El Paso, Texas, a U.S.-Mexico border city where its impact remains poorly understood. We employed wastewater analysis to detect xylazine and examine its potential correlation with fentanyl use over a 14-month period (June 2023 to July 2024). Our results show that xylazine was detected in wastewater samples from three of the four treatment plants serving the city. The prevalence of xylazine was heterogeneous, with the highest detection rate of 29% observed in one sewershed. All samples on xylazine-positive days also tested positive for norfentanyl, a fentanyl metabolite, demonstrating the widespread fentanyl consumption. Notably, sewersheds with higher xylazine detection exhibited significantly higher fentanyl loads, suggesting a community-level association between the two substances use. This study provides the first evidence of xylazine in Texas wastewater and highlights the urgent need for enhanced monitoring and targeted public health interventions to mitigate the growing threat of xylazine, particularly in border communities affected by the opioid crisis.

## 1. Introduction

Xylazine is a veterinary sedative and an emerging threat in the drug overdose crisis (Habib et al., 2024; Solanki et al., 2024). Its presence in overdose deaths has grown steadily since its first identification as an adulterant in Puerto Rico in the early 2000s. Xylazine (known as “tranq” in street drugs) is often found mixed with fentanyl and other drugs (Gupta et al., 2023; Johnson et al., 2021; Torruella, 2011; Zhu, 2023). The addition of xylazine extends the effects of opioids, while also increasing the risk of fatal overdose (D’Orazio et al., 2023; Papudesi et al., 2024). Xylazine is not an opioid. The treatment of Narcan/Naloxone against fentanyl is ineffective against xylazine (CDC, 2024; Zagorski et al., 2023). Recognizing this threat, the White House Office of National Drug Control Policy (ONDCP) declared xylazine as an emerging threat in April 2023 (The White House, 2023).

There are many challenges in addressing the xylazine threat. First, there is limited data on its prevalence and distribution, as xylazine is not routinely included in standard toxicology screens (Kariisa, 2021; Silva-Torres and Mozayani, 2024; Thangada, 2021). This leads to underreporting and an incomplete understanding of its true impact. Second, traditional surveillance methods, such as surveys and clinical reports, often suffer from delays of months or even years (CDC-National Center for Health Statistics, 2024; Spencer, 2016; Keshaviah et al., 2021), which hinders rapid response. Third, the illicit nature of xylazine use makes it difficult to accurately assess consumption patterns through conventional approaches (Ayub et al., 2023a). Finally, the varying legal status of xylazine across jurisdictions complicates coordinated monitoring and intervention efforts. Currently, only Florida has explicitly banned xylazine, while it remains unregulated in many other states (Cano et al., 2023; Drug Enforcement Administration, 2022; Sara, 2023). These challenges impede timely and effective public health responses to address the xylazine threat within the broader opioid crisis.

Wastewater-based epidemiology (WBE) offers a promising approach to overcoming many challenges in tracking drug use at the community level (Ahmed et al., 2023; Fontanals et al., 2024; Kirby, 2021; Sridhar et al., 2022; Wright and Adhikari, 2023a). Innovative methods have analyzed wastewater to detect and quantify drugs and their metabolites, providing near real-time data on drug use trends (Delcher et al., 2024; Luo et al., 2023). Utilizing excretion rates and wastewater flow data, wastewater data can be used to estimate drug consumption levels in the community or ‘sewershed’. The noninvasive nature of wastewater sampling also allows for continuous monitoring without the delays associated with traditional surveillance methods (Diamond et al., 2022). Researchers have successfully applied WBE to monitor the use of opioids, stimulants, and other drugs nationwide (Bishop et al., 2020; European Union Drugs Agency, 2024; Gerrity et al., 2011; Lin et al., 2021; Sulej-Suchomska et al., 2020; Wright and Adhikari, 2023b). A two-year wastewater surveillance program in rural Massachusetts revealed 100% detection frequency for ten opioids and stimulant drugs (Luo et al., 2023). A recent study in Kentucky detected xylazine in wastewater samples across the state (Delcher et al., 2024). These studies demonstrate the potential of WBE in providing timely insights into emerging drug trends and complementing existing health monitoring programs.

Our study focuses on El Paso, Texas, the sixth largest city in the state and a US-Mexico border city known for its major trade hub. El Paso County’s overdose death rate nearly doubled from 11.3 to 21.8 per 100,000 residents between 2018-2019 and 2022-2023, with 58% of these deaths involving fentanyl (The Houston Chronicle, 2024). The prevalence of xylazine use in El Paso is unclear, but the city is located within a region of concern based on DEA seizure reports indicating xylazine increasingly found mixed into fentanyl (Briano, 2023; Rios, 2023). Furthermore, a DEA Joint Intelligence Report showed a staggering 1127% increase in deaths involving xylazine across the southern United States (Drug Enforcement Administration, 2022). Giving these concerning trends, the aim of this study is to detect the presence of xylazine and examine its potential correlation with fentanyl use in El Paso’s wastewater over 14 months (June 2023 to July 2024, **Fig. 1**). This investigation is critical for understanding the evolving drug trends in this important border community. To our knowledge, no similar analysis has been reported in Texas.

**Figure 1.**
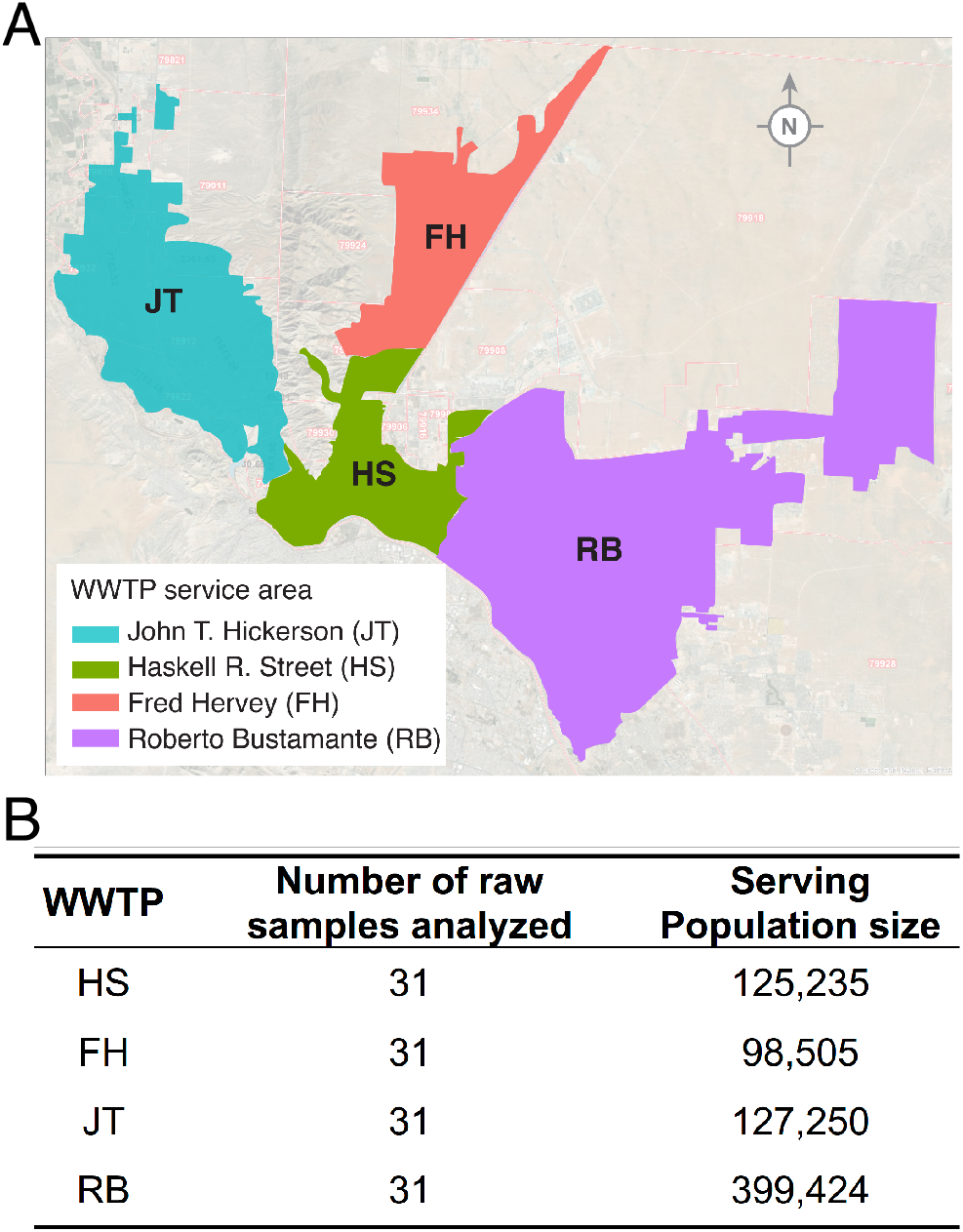
The map and sampling information for El Paso, Texas. (A) Map showing the locations of wastewater treatment plants (WWTPs) in El Paso, Texas. (B) Wastewater samples analyzed from each WWTP, along with the corresponding population size served by each plant.

## 2. Materials and Methods

### 2.1 Wastewater sample collection

The 24-hr composite, raw influent wastewater samples were collected from the four wastewater treatment plants (WWTPs) in the city of El Paso, Texas, from June 26, 2023, to July 15, 2024. The four WWTPs including Fred Hervey (FH), Haskell R. Street (HS), John T. Hickerson (JT), and Roberto Bustamante (RB), serves ∼751,982 customers in total in the city (**Fig. 1A**) (Oghuan et al., 2023). Samples were shipped overnight on ice to Houston for analysis. The samples were stored at -20°C and accumulated for analysis. A total of 124 samples (31 from each WWTP) were collected and tested in this study (**Fig. 1B**).

### 2.2 Sample Preparation and Quantification with LC-MS/MS

Raw wastewater samples (35 mL) were centrifuged, transferred, and further filtered with 0.22 um polyethersulfone membrane (Steriflip, Millipore Sigma) to remove remaining detritus. The filtrate (30 mL) was extracted using solid-phase extraction cartridges on an extraction manifold (Waters, USA). This approach allowed for accurate quantification by accounting for variability in sample processing. Samples were acidified to acidified to pH 2.5 using hydrochloric acid (HCl, Millipore Sigma) and then loaded onto an Oasis MCX cartridge (Waters), which was pre-conditioned sequentially with HPLC-grade methanol (Millipore Sigma), HPLC-grade water (Millipore Sigma), and 0.1% HCl in HPLC water (v/v). A vacuum pump was applied. The sample bottles were sequentially rinsed with water and 0.1% v/v HCl, and the rinse solutions were also loaded onto the cartridge.

Methanol was used to wash the cartridges, and the vacuum was applied for 30 seconds to eliminate any residual wash solvent. The cartridge was then air-dried at room temperature. The analytes were eluted from the cartridge using a 2 mL mixed solution of tert-butyl methyl ether, isopropanol, and ammonium hydroxide at a ratio of 78:20:2 (v/v/v, Millipore Sigma). The vacuum was again applied for 30 seconds to flush out any residual elution buffer. The eluent was collected in 15 mL tubes, dried with a nitrogen gas stream, reconstituted with 150 μL of 50% methanol centrifuged at 15,000 relative centrifugal field (rcf) for 15 minutes. The supernatants were transferred to sample vials and 3 μL was injected for liquid chromatography tandem mass spectrometry (LC-MS/MS) analysis using the Thermo TSQ Quantis coupled with a Thermo Vanquish UHPLC (Thermo Fisher Scientific, CA). The analytes were separated on an Agilent XDB-C18 column (4.6 mm × 50 mm, 3 μm), and eluted by a water-acetonitrile mobile phase system (both containing 0.1% formic acid, v/v) increasing from 15% to 95% organic phase within a 6-min run. The flow rate was set at 0.3 mL/min. The column temperature was set at 40 °C. Analytes and IS were monitored under the selected reaction monitoring mode coupled with a positive electrospray ionization source.

To establish and validate the protocol, controls of four different concentrations were prepared for each experimental batch by spiking HPLC-grade water (Millipore Sigma) with certified reference xylazine and deuterated xylazine (Cayman Chemical, Ann Arbor, MI) at concentrations of 0.1 ng/L, 1 ng/L, 10 ng/L, and 100 ng/L. Xylazine was consistently detected at the 0.1 ng/L level, but not at lower concentrations (0.01 ng/L), so the limit of detection to be determined as 0.1 ng/L. A deuterated xylazine-d6 (analytical reference standard, Cayman Chemical) was also added into each wastewater sample at a concentration of 100 ng/L before starting the extraction protocol.

The concentration of xylazine in each sample was quantified using two methods: a calibration curve and an internal deuterated xylazine (xylazine-d6). For samples collected on days when xylazine was detected, we also analyzed norfentanyl, a human metabolite of fentanyl. Norfentanyl quantification was performed using a standard reference (Cayman Chemical) for building a standard calibration curve following the same procedure.

### 2.3. Calculation of xylazine mass load and per-capita consumption

The xylazine mass load was calculated by multiplying the measured concentrations (ng/L) by the daily influent flow volume (Equation 1) (Bishop et al., 2020; Fontanals et al., 2024; Gushgari et al., 2019). Daily wastewater flow data (million gallons per day, MGD) were provided by the wastewater treatment plants. One MGD equals 3,785,412 liters. The per-capita consumption was then determined by dividing the xylazine mass load by the population size in each sewershed (Equation 2). This estimation does not account for the drug excretion rate into urine and stool, which is approximately 70% for xylazine based on studies conducted in rats (Veilleux-Lemieux et al., 2013), and 91% for fentanyl in human (Gushgari et al., 2019; Labroo et al., 1997). To date, no corresponding human excretion data for xylazine has been reported.

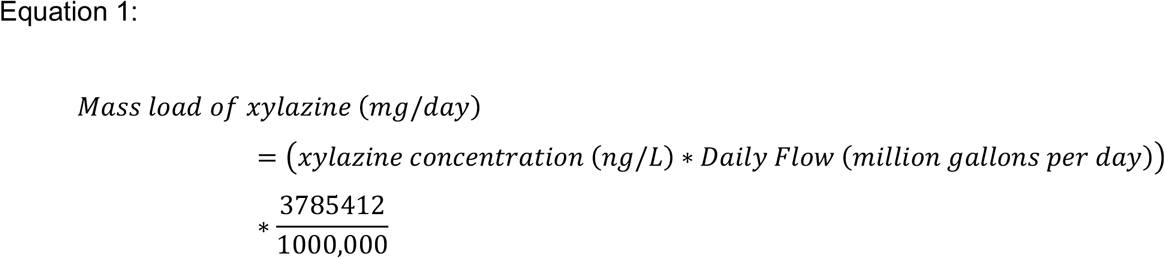

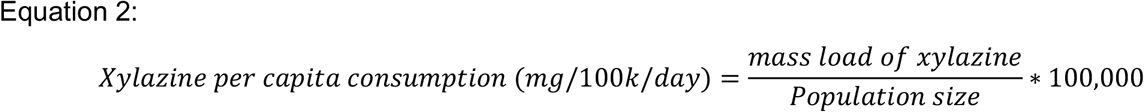

### 2.4. Statistical analysis

The Welch two-sample t-test was employed to assess differences in norfentanyl per-capita consumption loads between sewersheds (HS vs. JT and HS vs. RB). This test was chosen for its robustness in comparing groups with potentially unequal variances. A significance level of α = 0.05 was established, with p-values below this threshold indicating statistically significant differences between compared sewersheds. All statistical analyses were performed using R software (version 4.3.1), with the ‘t.test()’ function for the Welch t-test.

### 2.5. Data about Mortality-involving Xylazine from NFLIS Reports

Xylazine-involved mortality data was obtained from The National Forensic Laboratory Information System (NFLIS) (U.S. Drug Enforcement Administration, Division Control Division, 2024). NFLIS is a program under the supervision of the Drug Enforcement Administration (DEA) which serves as a repository for results of drug identification tests and related information submitted by participating forensic laboratories at the local, state, and federal levels. NFLIS data was queried to investigate reported xylazine-involved mortalities. We queried data from reports involving xylazine from 2019 to 2023 from the NFLIS Public Data Query System (DQS). The search term “xylazine” was applied for those years and the numbers of related drug reports were collected at the state (Texas) and national levels. The queried data was exported for analysis.

## 3. Results

### 3.1 Xylazine quantification methods and comparison

To ensure accurate quantification of xylazine in wastewater samples, we evaluated two analytical approaches: the standard calibration curve method and the internal deuterated standard method. **Fig. 2A** shows the standard calibration curve for xylazine quantification using LC-MS/MS analysis (Centazzo et al., 2019; Cruz-Cruz et al., 2021; Gushgari et al., 2019). The graph plots the peak area ratio of xylazine/xylazine-d6 against known spiking concentrations ranging from 0.1 to 100 ng/L, with 0.1 ng/L established as the limit of detection. A linear regression model applied to these data yielded a high coefficient of determination (R^2^ =0.996), demonstrating the method’s precision and linearity across a wide dynamic range. This robust calibration enables accurate determination of xylazine concentrations in unknown wastewater samples. In parallel, we employed an internal standard method by spiking each sample with deuterated xylazine (xylazine-d6). This approach allows for direct quantification by comparing the signal of the native xylazine to that of the deuterated standard, accounting for potential matrix effects and variations in instrument response. **Fig. 2B** shows the comparative analysis of xylazine quantification in positive wastewater samples using both methods. The close clustering of data points around the y=x line indicates strong correlation between the two methods, suggesting no significant difference in their performance for xylazine quantification in wastewater.

**Figure 2.**
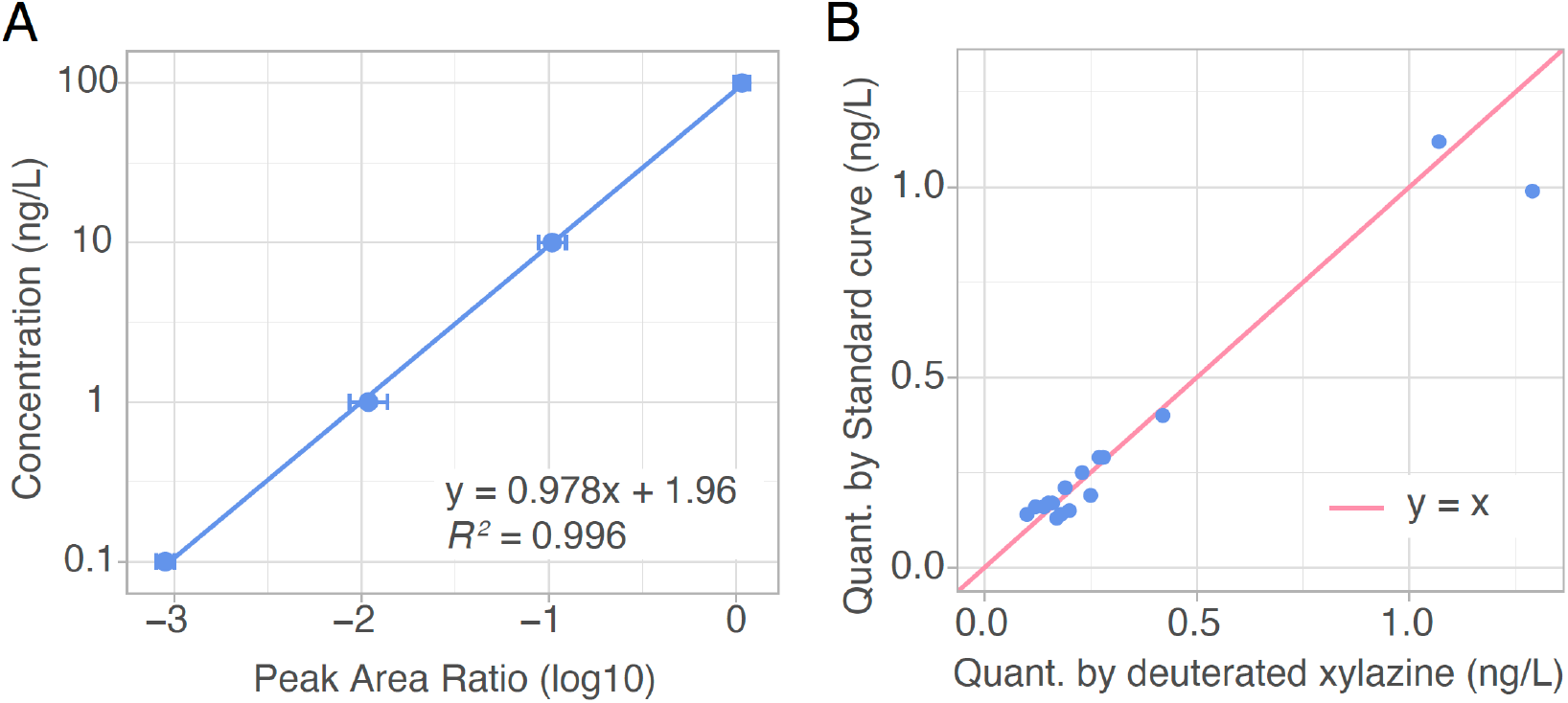
Compare the quantification methods of xylazine. **(A)** Standard calibration curve for xylazine quantification. The graph shows the relationship between the peak area ratio of xylazine and known spiking concentrations of the standards. Data points represent the mean of three independent batches (N=3), with error bars indicating standard deviation. **(B)** Comparison of xylazine concentrations using standard curve (y-axis) and internal deuterated xylazine (xylazine-d6).

### 3.2. Xylazine detection and heterogeneous prevalence across sewersheds

We analyzed 124 wastewater samples for xylazine from four wastewater treatment plants (**Fig. 3A**) serving the entire border city of El Paso, Texas, over a 14-month period from June 2023 to July 2024. LC-MS/MS results showed heterogeneous prevalence across sewersheds. Specifically, xylazine was detected in samples from 3 out of 4 WWTPs (FH, HS, and JT), with the highest positive detection rate found in the HS sewershed (29%, **Fig. 3B**). Notably, none of the samples from the RB WWTP tested positive for xylazine. The concentrations ranged from 0.1 ng/L (limit of detection) to 1.44 ng/L, quantified using deuterated xylazine (xylazine-d6) as an internal standard. To estimate xylazine usage rates, we incorporated data on wastewater flow volume and the population size served by each sewershed. While mass per-capita loads of xylazine remained relatively stable over time, notable peaks were observed in HS (41.3 mg/100k/day) and FH (28.8 mg/100k/day) in late February 2024. These results suggest varying patterns of xylazine use across different areas of El Paso, with some sewersheds showing higher and more frequent detection than others. The temporal stability in most areas, punctuated by occasional spikes, may indicate a consistent user base with periodic influxes of the drug or changes in usage patterns.

**Figure 3.**
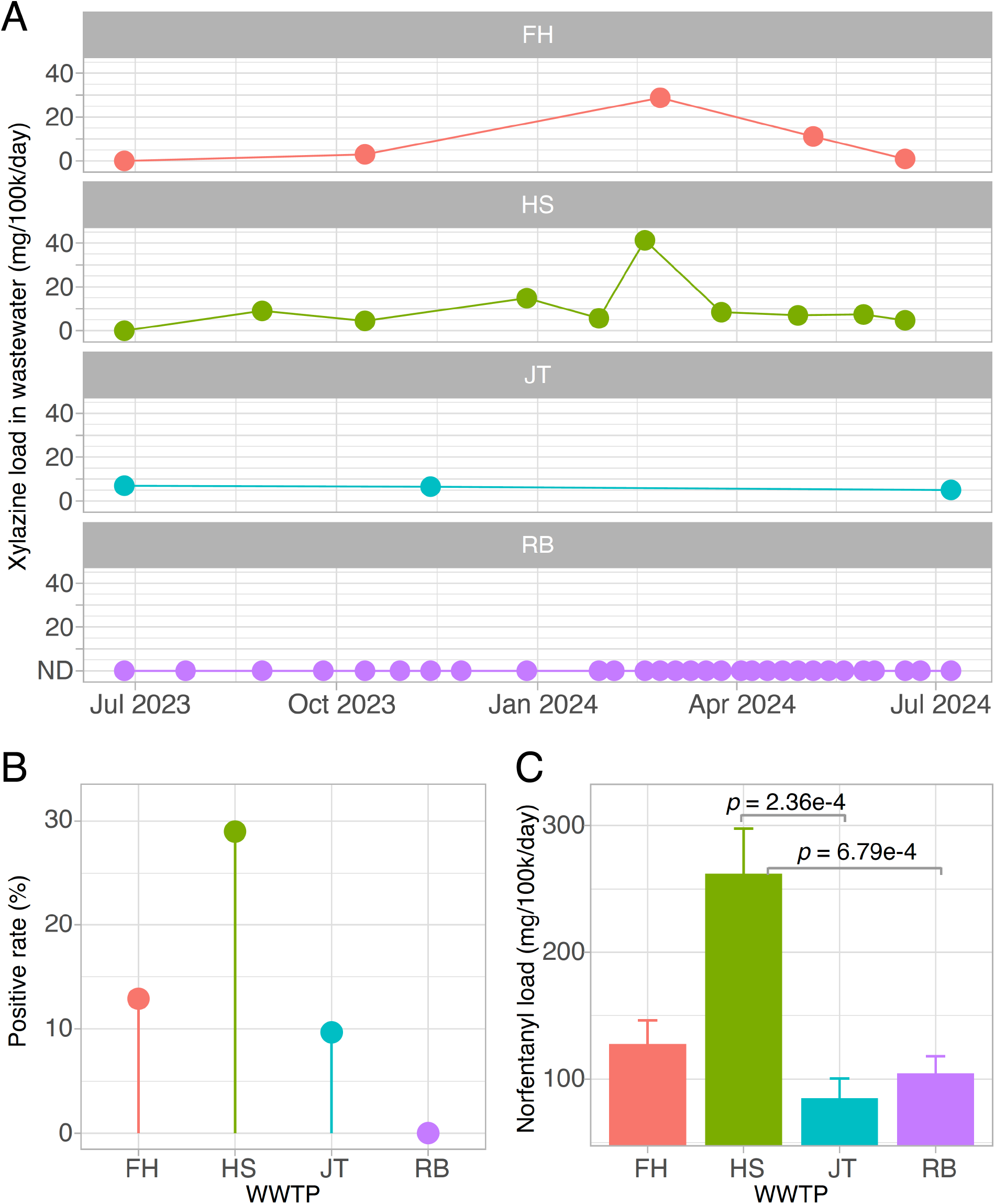
Xylazine detection in wastewater and its association with fentanyl use. (A) Temporal trends of xylazine per-capita load across the four sewersheds (FH, HS, JT, RB) in El Paso, Texas. All samples in RB were below the limit of detection (Points below the limit of detection for samples from FH, HS, and JT were omitted for visualization). (B) Prevalence (Positive detection rate) of xylazine in wastewater by sewershed. (C) Norfentanyl per-capita load (a metabolite of fentanyl consumption) across sewersheds. Xylazine and norfentanyl loads were calculated by multiplying the measured concentrations by the wastewater flow volume and normalized by the population size. P-value was obtained via the Welch two sample t-test.

### 3.3 Xylazine prevalence is associated with fentanyl consumption rate

To examine potential correlations between xylazine and opioid use, we tested for norfentanyl in all samples collected on days when xylazine was detected. This approach allowed us to compare norfentanyl loads across WWTPs on xylazine-positive days and investigate potential associations. In total, 56 samples from days with at least one xylazine-positive result across all sewersheds were analyzed. All samples tested positive for norfentanyl, with concentrations of 5.12±0.40 ng/L (mean±s.e.m.). This ubiquitous presence of norfentanyl suggests a high prevalence of fentanyl use across El Paso.

To further investigate potential relationships, we estimated and compared fentanyl consumption rates across sewersheds. The results showed that the HS sewershed had the highest fentanyl use, with a mass per-capita load of norfentanyl at 262.13±35.41 ng/L, followed by FH at 127.66±18.68 ng/L. Both HS and FH exhibited higher norfentanyl loads compared to JT (84.94±15.58 ng/L) and RB (104.53±13.46 ng/L) (**Fig. 3C**). Statistically significant differences were found between HS and JT, and between HS and RB. This pattern aligns with the higher prevalence of xylazine in HS and FH (**Fig. 3B**), highlighting a clear link between xylazine prevalence and fentanyl consumption burden in these areas of the city.

## 4. Discussion

This study provides the first evidence of xylazine presence in wastewater samples from El Paso, Texas, a major city on the U.S.-Mexico border. We detected xylazine in three out of four wastewater treatment plants serving the city, with the highest prevalence in the HS sewershed. Notably, we observed an association between xylazine prevalence and higher fentanyl consumption rates in the same areas, suggesting a potential link between the use of these substances. These findings provide a population-level perspective on the emerging xylazine threat in a region already grappling with the opioid crisis.

The detection of xylazine in El Paso wastewater aligns with the rising trend of xylazine-involved mortalities reported across the United States, particularly in the South. According to the DEA, the Southern U.S. had the highest increase in xylazine identifications and xylazine-involving overdose mortalities nationwide (Drug Enforcement Administration, 2022). This is consistent with the national trend of xylazine-involved mortalities based on data from the NFLIS (**Fig. 4**). In Texas, while xylazine-involving deaths started a decrease in 2021, the total mortality in 2023 was approximately nine-fold higher than in 2019 (U.S. Drug Enforcement Administration, Division Control Division, 2024). Although public records have not yet indicated xylazine-related mortalities in El Paso, the DEA has reported seizing fentanyl and xylazine mixtures within the county (Briano, 2023). Our results provide direct evidence of xylazine’s presence in the city’s wastewater, potentially indicating a more widespread issue than previously recognized. The co-occurrence of higher fentanyl consumption levels in areas with more frequent xylazine detection is particularly concerning, because the combination of these drugs is known to drastically increase the risk of fatal overdoses (Ayub et al., 2023b; Montero et al., 2022), and the standard opioid antagonists like naloxone are ineffective against its sedative effects (Bedard et al., 2024; Gupta et al., 2023).

**Figure 4.**
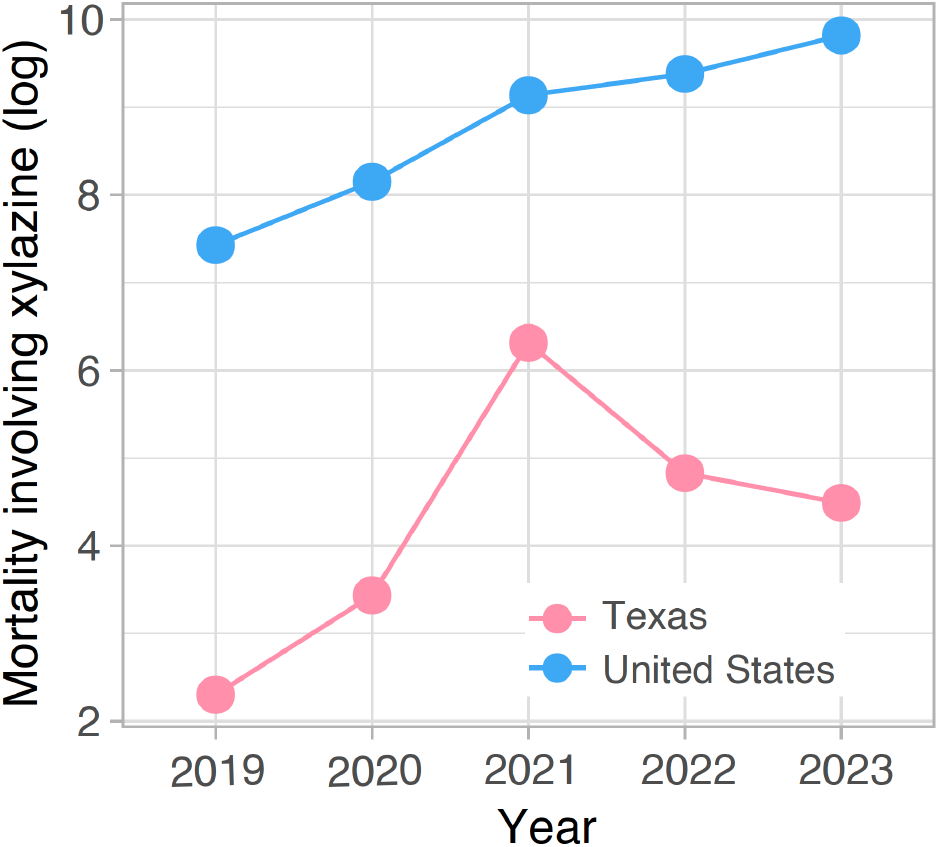
Xylazine-involved mortality trends in Texas and the United States from 2019 to 2023. Original data were obtained from the NFLIS data query system (y-axis: natural logarithm).

This study has three limitations that should be considered when interpreting the results. First, in estimating xylazine consumption rates, we did not account for the excretion rate (approximately 70% in rats, unknown in humans), and potential losses in the sewage system (Veilleux-Lemieux et al., 2013). Including these factors would likely increase our consumption rate estimates and allow for more accurate dose estimations per sewershed. Second, while xylazine detection likely indicates human use, especially given its correlation with fentanyl levels, we cannot entirely rule out veterinary sources. However, many registered veterinary clinics are in the RB sewershed, where no xylazine was detected, and many animal ranches are in the JT sewershed (El Paso Veterinary Medical Association, 2024). The absence of local xylazine-involved case data makes it challenging to correlate our findings with clinical outcomes. Finally, wastewater data may not exactly reflect localized use patterns due to population movement within the city and daily influx of visitors. El Paso receives an average of 15,532 pedestrians, 35,712 passenger vehicles, and 2,937 commercial vehicles daily through its ports of entry (The International Bridges Steering Committee, n.d.). These limitations emphasize the need for future research to differentiate between human and animal sources of xylazine, and correlate wastewater findings with clinical data.

Despite these limitations, our study represents the first demonstration of xylazine presence in Texas wastewater and reveals a heterogeneous prevalence across sewersheds and a clear association with elevated fentanyl use. The findings highlight the need for continued surveillance and targeted public health interventions in El Paso and similar communities experiencing an elevated drug overdose burden. Proactive, data-driven strategies informed by wastewater analysis can help guide the allocation of resources and implementation of harm reduction measures. Additionally, this approach can serve as a tool to evaluate the effectiveness of policies and interventions aimed at addressing the xylazine and opioid crisis (Sugarman et al., 2024). By tracking changes in drug use patterns over time, communities can assess the impact of their efforts and make data-driven adjustments to their strategies.

## Data Availability

Data and Code Availability
Data and code in this study will be shared with the paper publication.

## Data and Code Availability

Data in this study will be shared with the paper publication. Scripts will be shared on GitHub.

## Acknowledgments

This work is supported by Southwest Center for Occupational and Environmental Health (SWCOEH), the Centers for Disease Control and Prevention (CDC) / National Institute for Occupational Safety and Health (NIOSH) Education and Research Center (T42OH008421) at The University of Texas Health Science Center at Houston School of Public Health. The authors are also grateful for the support by National Institute of Drug Abuse (R01DA059394-01A1). We acknowledge the NMR and Drug Metabolism Core at Baylor College of Medicine for the drug quantification. KMJ is supported by the Texas Epidemic Public Health Institute. We are also grateful to El Paso Water for providing the samples.

